# Evaluating the generalisability of region-naïve machine learning algorithms for the identification of epilepsy in low-resource settings

**DOI:** 10.1101/2024.03.25.24304872

**Authors:** Ioana Duta, Symon M Kariuki, Anthony K Ngugi, Angelina Kakooza Mwesige, Honorati Masanja, Seth Owusu-Agyei, Ryan Wagner, J Helen Cross, Josemir W Sander, Charles R. Newton, Arjune Sen, Gabriel Davis Jones

## Abstract

**Objectives:** Approximately 80% of people with epilepsy live in low- and middle-income countries (LMICs), where limited resources and stigma hinder accurate diagnosis and treatment. Clinical machine learning models have demonstrated substantial promise in supporting the diagnostic process in LMICs without relying on specialised or trained personnel. How well these models generalise to naïve regions is, however, underexplored. Here, we use a novel approach to assess the suitability and applicability of such clinical tools for diagnosing active convulsive epilepsy in settings beyond their original training contexts.

**Methods:** We sourced data from the Study of Epidemiology of Epilepsy in Demographic Sites dataset, which includes demographic information and clinical variables related to diagnosing epilepsy across five sub-Saharan African sites. For each site, we developed a region-specific (single-site) predictive model for epilepsy and evaluated its performance on other sites. We then iteratively added sites to a multi-site model and evaluated its performance on the omitted regions. Model performances and parameters were then compared across every permutation of sites. We used a leave-one-site-out cross-validation analysis to assess the impact of incorporating individual site data in the model.

**Results:** Single-site clinical models performed well within their own regions, but worse in general when evaluated on other regions (p<0.05). Model weights and optimal thresholds varied markedly across sites. When the models were trained using data from an increasing number of sites, mean internal performance decreased while external performance improved.

**Conclusions:** Clinical models for epilepsy diagnosis in LMICs demonstrate characteristic traits of ML models, such as limited generalisability and a trade-off between internal and external performance. The relationship between predictors and model outcomes also varies across sites, suggesting the need to update specific aspects of the model with local data before broader implementation. Variations are likely to be specific to the cultural context of diagnosis. We recommend developing models adapted to the cultures and contexts of their intended deployment and caution against deploying region- and culture-naïve models without thorough prior evaluation.

**Key points:**

- Machine learning-driven clinical tools are becoming more prevalent in low-resource settings; however, their general performance across regions is not fully established. Given their potential impact, it is crucial models are robust, safe and appropriately deployed
- Models perform poorly when making predictions for regions that were not included in their training data, as opposed to sites that were
- Models trained on different regions can have different optimal parameters and thresholds for performance in practice
- There is a trade-off between internal and external performance, where a model with better external performance usually has worse internal performance but is generally more robust overall

**SEEDS collaborators:**

- Agincourt HDSS, South Africa: Ryan Wagner, Rhian Twine, Myles Connor, F. Xavier Gómez-Olivé, Mark Collinson (and INDEPTH Network, Accra, Ghana), Kathleen Kahn (and INDEPTH Network, Accra, Ghana), Stephen Tollman (and INDEPTH Network, Accra, Ghana)
- Ifakara HDSS, Tanzania: Honratio Masanja (and INDEPTH Network, Accra, Ghana), Alexander Mathew
- Iganga/Mayuge HDSS, Uganda: Angelina Kakooza, George Pariyo, Stefan Peterson (and Uppsala University, Dept of Women’s and Children’s Health, IMCH; Karolinska Institutet, Div. of Global Health, IHCAR; Makerere University School of Public Health), Donald Ndyomughenyi
- Kilifi HDSS, Kenya: Anthony K Ngugi, Rachael Odhiambo, Eddie Chengo, Martin Chabi, Evasius Bauni, Gathoni Kamuyu, Victor Mung’ala Odera, James O Mageto, Isaac Egesa, Clarah Khalayi, Charles R Newton
- Kintampo HDSS, Ghana: Ken Ae-Ngibise, Bright Akpalu, Albert Akpalu, Francic Agbokey, Patrick Adjei, Seth Owusu-Agyei, Victor Duko (and INDEPTH Network, Accra, Ghana)
- London School of Hygiene and Tropical Medicine: Christian Bottomley, Immo Kleinschmidt
- Institute of Psychiatry, King’s College London: Victor CK Doku
- UCL Queen Square Institute of Neurology, London: Josemir W Sander
- Swiss Tropical Institute: Peter Odermatt

## Introduction

Epilepsy is a common neurological condition that disproportionately affects people from disadvantaged socio-economic groups. Studies estimate that up to 75 million people have epilepsy worldwide, with approximately 80% in low- and middle-income countries (LMICs)(1). Epilepsy accounts for over 13 million disability-adjusted life years annually and over 0.5% of the global burden of disease. While an estimated 70% of people with epilepsy could live seizure-free with anti-seizure medications, over 75% of those living with epilepsy in LMICs cannot obtain an appropriate diagnosis or any treatment (2)(3).

The diagnosis of epilepsy requires training, skilled personnel, time and additional resources that are scarce in LMICs, for example access to specialised equipment such as electroencephalograms (EEGs) (4). A trained clinician’s expertise is irreplaceable, but training costs and retention of skilled personnel can impede access to diagnosis in LMICs. In such settings, diagnostic tools that require less expertise, experience or specialist training could empower primary healthcare workers to triage and prioritise people who may have epilepsy for referral (5).

Clinical machine learning (ML) models therefore offer a practical solution for epilepsy diagnosis in low-resource settings. Such models have demonstrated promising outcomes for epilepsy diagnosis (6) and treatment (7). Thus, their application in LMICs could reduce the diagnostic and treatment gaps (8). Given the potential impact of such models, their relevance, robustness and appropriate deployment are crucial.

ML models developed on data from one region are not inherently reliable for use elsewhere without prior validation. A measure of model robustness in new settings is termed ‘generalisability’. Failure of a model to generalise sufficiently to a novel setting can be due to, for example, differences in clinical phenomenology or patient self-reporting across the regions. This is particularly applicable in epilepsy, where clinical diagnosis can be nuanced (9). This phenomenon can also be attributed to the model ‘overfitting’, where the model learns to make predictions based on biases in the dataset rather than developing a robust method for delineating between the desired diagnoses of interest (or absence thereof) (10). In the case of diagnostic tools, this can have substantial consequences for the population on whom the model is deployed, resulting in missed cases, over-diagnosis, wasted resources and even mistrust of the technology (10). The issue of generalizability has been well described in other medical domains (10).

We have previously developed a predictive model to support epilepsy diagnosis in LMICs (6), trained on data from five sub-Saharan African regions. This study aims to describe the necessity of comprehensive cross-regional validation. We investigate the suitability of diagnostic models for deployment in regions which do not contribute data to the models’ training. We also consider generalised seizures (9) separately due to their relatively homogeneous presentation, to investigate how incorporating seizure subtype in a model’s training influences the model’s performance both on sites that were included in training and those that were not.

## Methods

### Data acquisition, study design and pre-processing

We sourced data from the Study of Epidemiology of Epilepsy in Demographic Surveillance Sites (SEEDS), which assessed the prevalence and risk factors of active convulsive epilepsy (ACE) in five sites across sub-Saharan Africa: Agincourt (South Africa), Ifakara (Tanzania), Iganga (Uganda), Kilifi (Kenya) and Kintampo (Ghana). This dataset has been described in detail elsewhere. (11) The dataset comprised responses to an ACE-specific questionnaire. ACE was defined as two or more unprovoked seizures occurring at least 24 hours apart with at least one episode in the preceding year. Convulsive epilepsy was chosen explicitly because convulsions are more easily identified and are associated with higher morbidity, mortality and stigma. The protocol for the SEEDS study has been previously published (11). The outcome of this study was a clinical diagnosis of ACE (EEG supported where possible) confirmed by an epilepsy-specialised neurologist. Individuals with and without epilepsy (here considered as controls) are included in the dataset. Information on socio-demographic variables, historical risk factors and clinical history was collected for each participant. The resulting dataset comprised sociodemographic information and approximately 170 unique variables for each individual in five domains: clinical history, clinical examination, seizure description and EEG interpretation (6).

### Data pre-processing

We selected participants with either a confirmed diagnosis of ACE (cases) or absence of ACE (controls) following evaluation by a trained neurologist. We used predictors of epilepsy formatted as questions for the person with suspected epilepsy. These predictors have been reported and validated in previous studies from this cohort and were chosen for their maximally discriminative predictive ability to diagnose ACE (6). The predictors were:

1. During these episodes, have you ever bitten your tongue?
2. Have you ever wet yourself during these episodes?
3. During these episodes, do you lose contact with your surroundings?
4. Has anyone told you that you appear dazed during these episodes?
5. During these episodes, does your body stiffen?
6. Do you experience stomach-ache before these episodes?
7. Do you see odd things (e.g. flashes or bright lights) before these episodes occur?
8. Do you think anything brings on these episodes?

We retained only participants with a confirmed diagnosis (cases) or absence of ACE (controls). This was termed the “complete dataset”. We also considered a smaller subset in which only participants with either a confirmed “generalised” seizure type or absence of ACE were retained (referred to as the ‘generalised dataset’). The purpose was to minimise inherent heterogeneity in patient symptomatology due to multiple seizure types within a single individual. Selecting for generalised seizures reduces potential confounding across sites due to the inherently heterogeneous symptomatology of focal seizures and non-convulsive generalised epilepsy. The datasets were then separated by study site. Imputation and weighting were performed within each site separately. Predictors with missing values were imputed using Multiple Imputation by Chained Equation (12). To adjust for potential confounding between study sites due to participant sex, age and seizure type (general, focal, or ‘other’) we applied inverse propensity weighting (13).

Study site datasets were then split into model training and testing subsets. When a model was trained on data from the generalised dataset, it was tested on data from the generalised subset. The data processing procedure and the ten seizure/type site subsets and data processing are summarised in Table 1 and Supplementary Materials, Figure 1.

**Figure 1.**
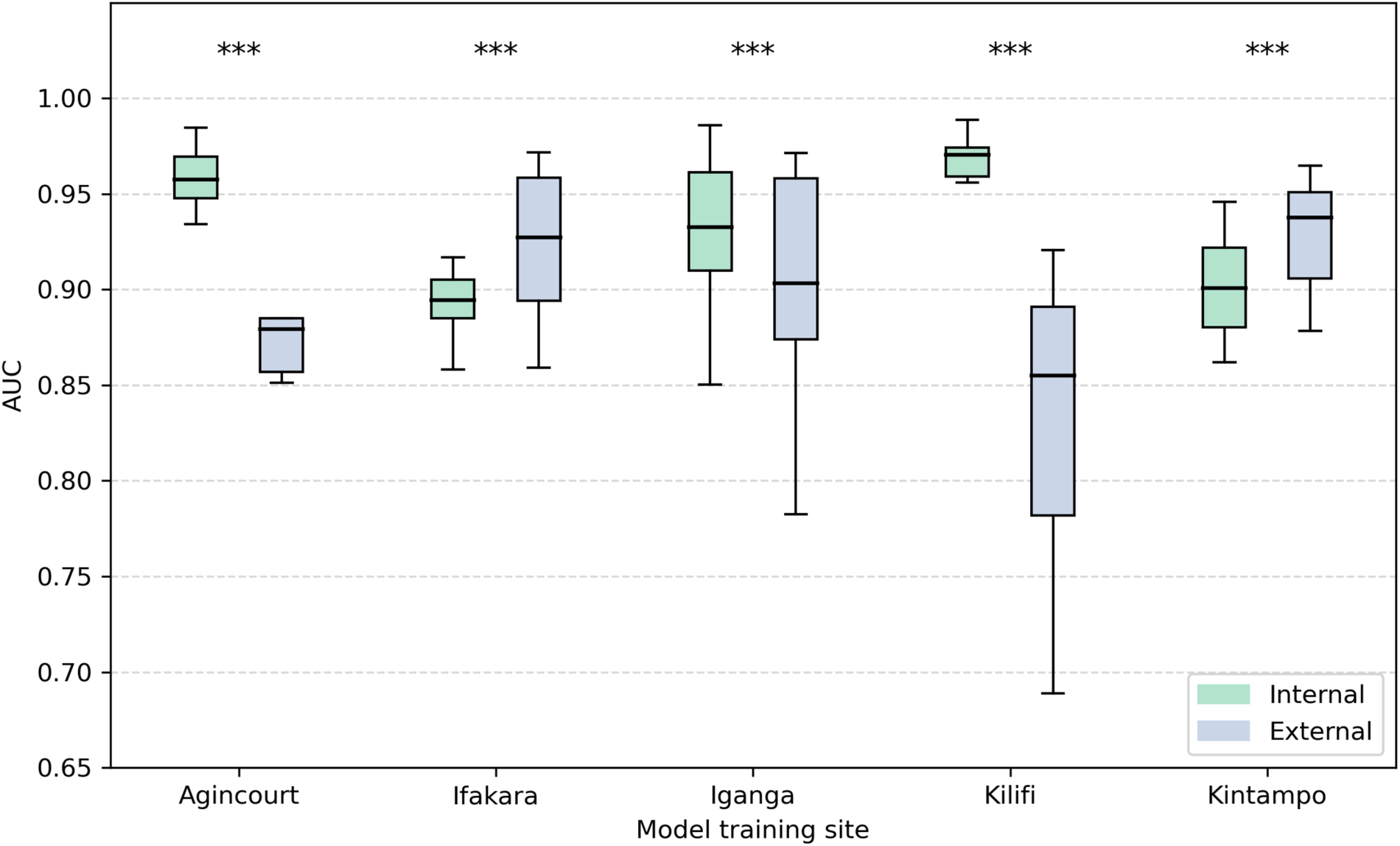
Comparative Internal and External AUC Performance Across Single-Site Models. The Area Under the Receiver Operating Characteristic Curve (AUC) performance of machine learning models trained and tested on data from single sites. Performance is compared between internal validations (training and testing on the same site) and external validations (training on one site and testing on others). Five distinct study sites in sub-Saharan Africa are evaluated: Agincourt (South Africa), Ifakara (Tanzania), Iganga (Uganda), Kilifi (Kenya), and Kintampo (Ghana). Boxplots describe the distribution of AUC values obtained through bootstrap resampling, indicating the variance within internal and external validations. At Agincourt, the internal AUC is 0.96 (interquartile range (IQR) 0.95– 0.97), while the external AUC is 0.88 (IQR 0.86–0.88), demonstrating a statistically significant difference with a decrease of approximately 0.08 in performance when models are externally validated. Similarly, Kilifi shows an internal AUC of 0.97 (IQR 0.96–0.97) against an external AUC of 0.84 (IQR 0.78–0.89), indicating a significant decline in external validation performance. Iganga’s internal and external AUCs are 0.93 (0.91–0.96) and 0.88 (IQR 0.87–0.96), displaying a smaller yet significant discrepancy. In contrast, Ifakara and Kintampo exhibit a converse trend, where external AUCs 0.92 (IQR 0.89–0.96) and 0.92 (IQR 0.91–0.95) slightly exceed their internal counterparts 0.89 (0.89–0.91) and 0.90 (0.88–0.92), although these differences are also statistically significant. These findings underscore the variability in model generalizability and the importance of external validation when assessing the robustness of predictive models in healthcare settings. *** = p-value <0.001.

**Table 1.** Dataset breakdown. Table showing the size of each subset of the data as split by diagnostic class, epilepsy type and site. The rightmost column represents a subset of the middle column. Participants with no clinical diagnostic information were excluded from the dataset. ACE = active convulsive epilepsy.

|  |  | Seizure types |  |  |
| --- | --- | --- | --- | --- |
|  |  | Controls | Confirmed ACE | Confirmed ACE & only generalised seizures |
| Site | Agincourt | 380 | 334 | 184 |
|  | Ifakara | 678 | 465 | 223 |
|  | Iganga | 498 | 259 | 152 |
|  | Kilifi | 793 | 791 | 209 |
|  | Kintampo | 516 | 394 | 233 |
| Total |  | 2865 | 2243 | 1001 |

### Model development

The models trained were Logistic Regression, Support Vector Machine (SVM) using a linear kernel and Naive Bayes assuming a Bernoulli distribution. These models were selected because they are computationally efficient, well-established in clinical prediction modelling in epilepsy, and because they yield interpretable results (6)(14). They also have varied approaches to decision boundaries (15). If there were an observed difference in the performance of these algorithms between study sites, it would likely be more attributable to the data than the model, i.e. an issue with generalisability.

The primary performance metric was the area under the receiver-operator characteristic curve (AUC) (26). AUC is independent of a classification threshold, eliminating the need to identify a threshold parameter and is less sensitive to data imbalance than other metrics, for example accuracy.

To ensure robustness, five-fold cross-validation was used to determine the performance of models on the data they were trained on (internal validation). In this process, training data are randomly split into five balanced subsets; four subsets are used to train the model and the remaining subset is used to test the model and calculate the AUC. This is repeated a further four times (five folds total), ensuring each subset is used for testing. The median of the five AUC values and the interquartile range are then used to determine the algorithm’s average performance (16).

Models trained on data within only one study site are ‘single-site’ models. A model for every permutation of the three algorithms, five sites and two seizure types was trained, resulting in 30 single-site models. Five-fold cross-validation AUC scores were obtained for each single-site model. Each single-site model was tested on each of the other four study sites, resulting in four external performance AUC scores. These were used to evaluate the performance of the models when tested on data from different regions. The Kolmogorov-Smirnov (K-S) test was used to assess whether there was a significant difference between the single-site AUC when tested on data from within the site (internal performance) and data from the other four study sites (external performance). The magnitudes, signs and ranges of the weights assigned to each predictor in each of the Logistic Regression models were also compared.

For each single-site model, we compared a set of incremental thresholds (20%, 50% and 80%) for classifying the probability output from each algorithm into a designation of ACE or control. For example, cases with a probability of less than 20% were assigned to controls and otherwise were assigned ACE. These thresholds were chosen to explore their effect on accuracy. The accuracies were calculated for the five study sites based on these classifications. We compared the relative change in accuracy between each study site from which a single-site model was developed with the accuracy from each separate study site. This analysis adds an additional dimension, as it extends the evaluation from AUC to a more clinical scenario where the direct classification of either ACE or control is important.

We then developed an iteratively inclusive multi-site model to evaluate the effect of adding additional study sites. Beginning with one randomly chosen study site, a model for each of the three algorithms was developed. The performance of each model was assessed on the remaining study sites. Another randomly chosen site was added to the model and the performance then re-calculated for the remaining unincluded study sites. This was repeated until every site except one (four training sites; one testing site) was included in the multi-site model, resulting in a leave-one-site-out (LOSO) model. We repeated this procedure until every permutation of study site orders of inclusion was achieved. Lastly, we developed a multi-site model that incorporated all study sites and split the data 70:30% into model training and validation subsets.

### Statistical Analysis

Continuous variables were binned. The two-sided Mann-Whitney test, with a significance level of 0.01, was used to evaluate for differences between internal and external datasets in the single-site models. The two-sample Kolmogorov-Smirnov (K-S) test, with a significance threshold of 0.05, was used to assess the significance of differences in model performances resulting from training data on different sites. Analysis was performed using Python 3 (17). Libraries used were Pandas (18), NumPy (19), SciPy (20), SKLearn (21), Seaborn (22), Pyplot (23) and Plotnine (24).

## Results

The complete dataset comprised 5,108 people with suspected epilepsy: 2,243 with confirmed ACE (44%) and 2,865 (56%) confirmed to not have epilepsy. Each participant was weighted such that there was a balance between the outcomes across sites and confounders. Missingness in the raw data appeared random (Supplementary Figure 2).

### Single-site models perform worse in new settings

Single-site models performed worse on average when tested on new sites. The median AUC when each single-site model was evaluated on novel data acquired from same site was 0.94 (IQR 0.90–0.96). However, when these models were tested with data from other study sites, the median AUC decreased significantly to 0.91 (IQR 0.89–0.93, p<0.01). Agincourt and Kilifi demonstrated the greatest difference between internal and external AUCs (Agincourt 0.96 internal, 0.88 external; p value <0.001, difference = -0.08 (7.9%); Kilifi 0.97 internal, 0.84 internal, difference = -0.13 (13%), p<0.001). Table 2 and Figure 1 show a comparison of the AUC values across each study site for internal and external validation.

**Table 2:** Single-site models perform worse in new settings.

| Site | Internal | External | Difference | P value |
| --- | --- | --- | --- | --- |
| Agincourt | 0.96<br>(0.95–0.97) | 0.88<br>(0.86–0.88) | -0.08<br>(7.9%) | <0.001 |
| Ifakara | 0.89<br>(0.89–0.91) | 0.92<br>(0.89–0.96) | 0.02<br>(2.7%) | <0.001 |
| Iganga | 0.93<br>(0.91–0.96) | 0.88<br>(0.87–0.96) | -0.05<br>(5.1%) | <0.001 |
| Kilifi | 0.97<br>(0.96–0.97) | 0.84<br>(0.78–0.89) | -0.13<br>(13.0%) | <0.001 |
| Kintampo | 0.90<br>(0.88–0.92) | 0.92<br>(0.91–0.95) | 0.03<br>(2.8%) | <0.001 |

Hypothesis tests comparing the performance scores resulting from the validation of a single-site model on a) the site whose data they were trained on and b) the other were statistically significant for eleven of thirty cases (see Table 1: ten data subsets, three models). There were seven significant differences for the dataset that considered all cases, and four for the generalised seizure dataset (Supplementary Materials, Table 1). Each site had six tests: of these, Agincourt and Kilifi each had five significant tests, while the other sites had at least five insignificant tests.

The weights assigned to the predictors of ACE in the logistic regression model differed between sites (Figure 2). All eight predictors’ weights had broad ranges, both when not accounting for seizure type (median range of weights 0.97, [IQR 0.72-1.5]) and when including only cases with generalised seizures (median range of weights 1.01 [IQR 0.86-1.3]). Apart from two variables, all variable weights were negative in some site models and positive in others. Only one predictor of ACE had consistently negative weights (corresponding to the question ‘Do you think anything brings on these episodes?’: range 0.31 when considering all controls, 0.75 for generalised seizures), and only one was consistently positive (corresponding to the question ‘Have you ever wet yourself during these episodes?’: range 0.72 when considering all controls, 0.49 for generalised seizures).

**Figure 2.**
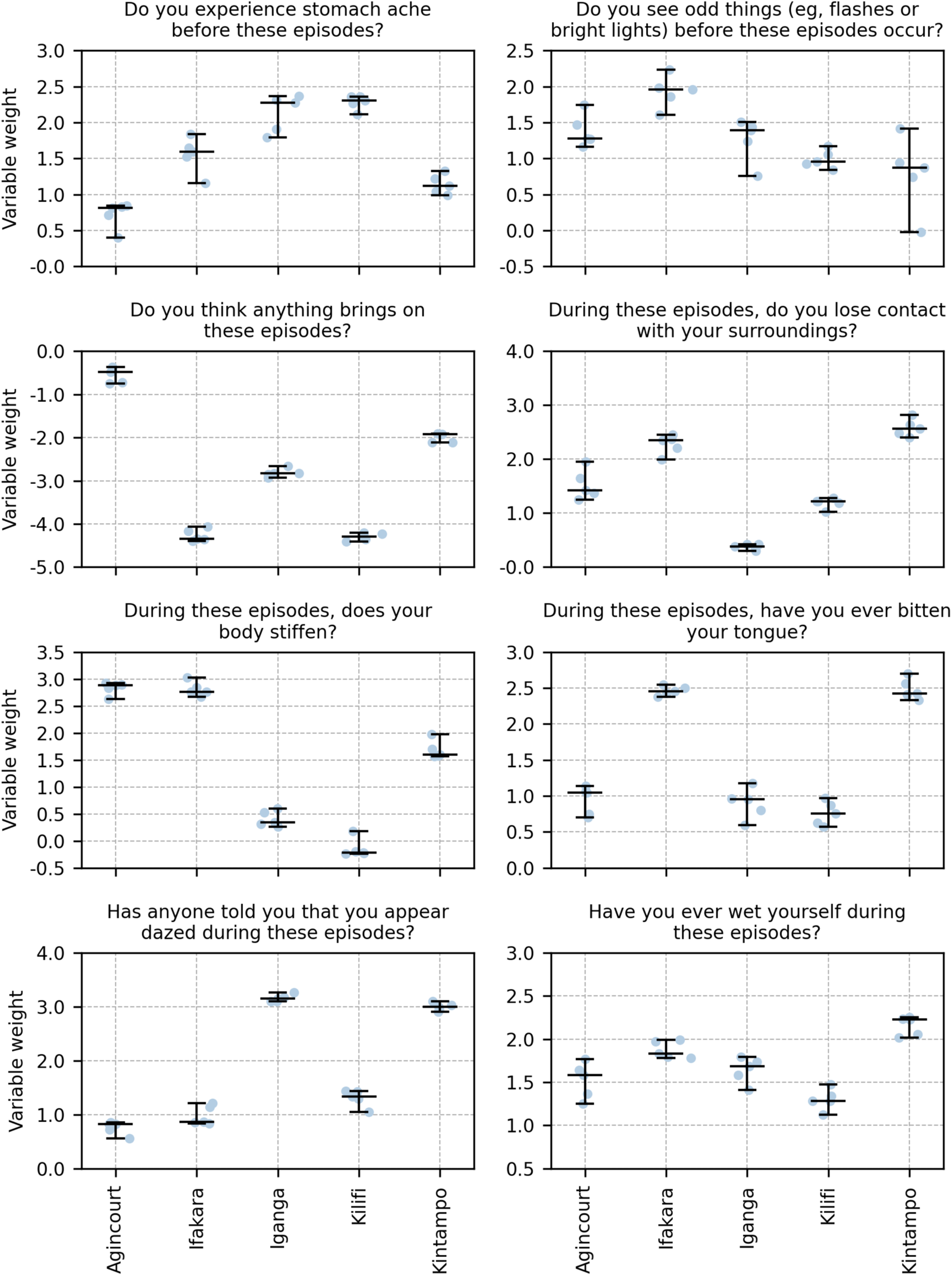
Variation of model weights between sites. Boxplot illustrating the values taken by the weights in each logistic regression model trained on a single site. Models were trained on a dataset in which positive cases were limited to participants with generalised epilepsy. All weights show a spread of values, and most change sign between sites. Only one covariate’s weights were consistently positive (‘Have you ever wet yourself during these episodes?’) and only one covariate’s weights were consistently negative (‘Do you think anything brings on these episodes?’). In this context positive weights correspond to a positive association with epilepsy, and negative weights to negative association. The horizontal line at the origin serves to clarify the threshold between positive and negative weights. Central line is median; lower edge of the box indicates first quartile, upper edge of the box indicates third quartile; points are extreme weight values.

Modulating the probability threshold for a positive diagnosis also resulted in variable accuracy. Increasing the threshold improved the performance in some sites and worsened performance in others. Figure 3 displays the difference in performance observed when models are tested outside of the development site. In Agincourt and Iganga, increasing the threshold from 0.2 to 0.5 to 0.8 resulted in a decrease in relative accuracy (Agincourt median -0.10, -0.13, -0.17, p<0.01; Iganga median -0.06, -0.18, - 0.19, p<0.01), while the inverse was true of Kintampo and Kilifi (Kilifi median -0.38, - 0.35, -0.05, p<0.01; Kintampo median -0.01, -0.01, 0.06, p<0.05).

**Figure 3.**
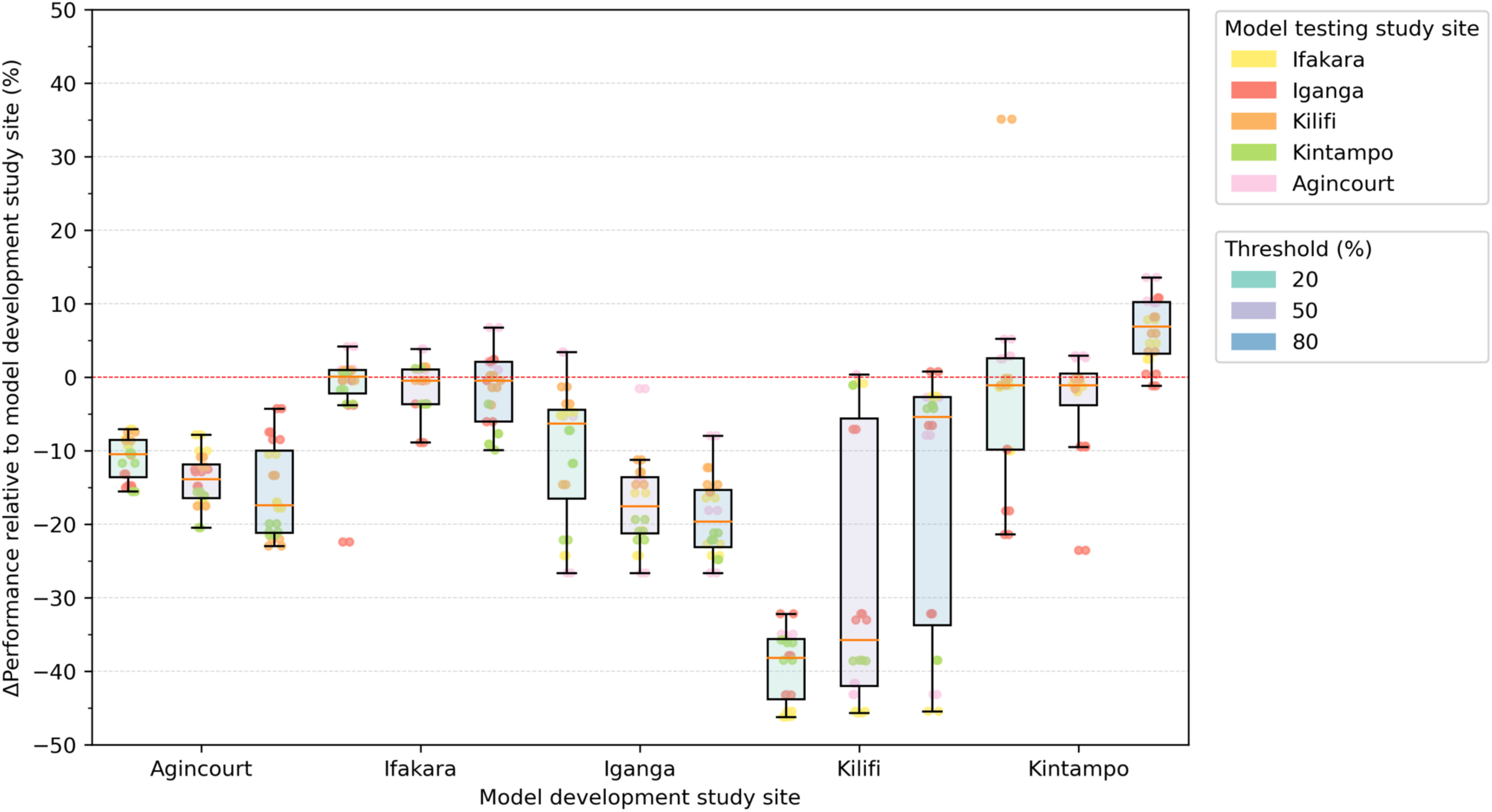
Effect of changing thresholds on accuracy. Heatmap showing how the accuracy of the logistic regression models was affected by changing the threshold from 0.2 to 0.8, for each one-site model. Models were trained on only one site’s data and tested on each of the other sites in turn. The performance of some site models worsened both internally and externally while the other sites’ performance improved. The overall mean change in accuracy was 10% (min 0.4%, max 28%, standard deviation 7.6%).

### Incremental site inclusion

As additional sites were incorporated into the model, internal performance declined (initial median AUC 0.94, IQR 0.11; final median AUC 0.93, IQR 0.01; p=0.06), and external performance improved (initial median 0.90, IQR 0.04; final median 0.92, IQR 0.02; p<0.01; Figure 5). At the initial stage of this process, when only one site was included in the training data, internal performance was greater than external (mean AUC difference 0.050, p<0.01) and all external performances were lower than all the internal performances.

At the final stage, the two measures converged to a small final difference, and all the data points from the external validation were higher than all the internal points (Figure 5). The mean difference between final performance scores was 0.001 (p-value 0.83).

### Leave One Site Out

Internal performance was higher in 3 sites: Ifakara (internal median 0.93, IQR 0.00; external median 0.92, IQR 0.06; p=0.55), Iganga (internal median 0.94, IQR 0.02; external median 0.91, IQR 0.03; p<0.01) and Kintampo (internal median 0.937, IQR 0.019; external median 0.89, IQR 0.006; p<0.01) (Figure 4). Kintampo’s external scores had a smaller range (p<0.01) while the other two sites had a larger external range (Ifakara: p<0.01; Iganga: p=0.80).

**Figure 4.**
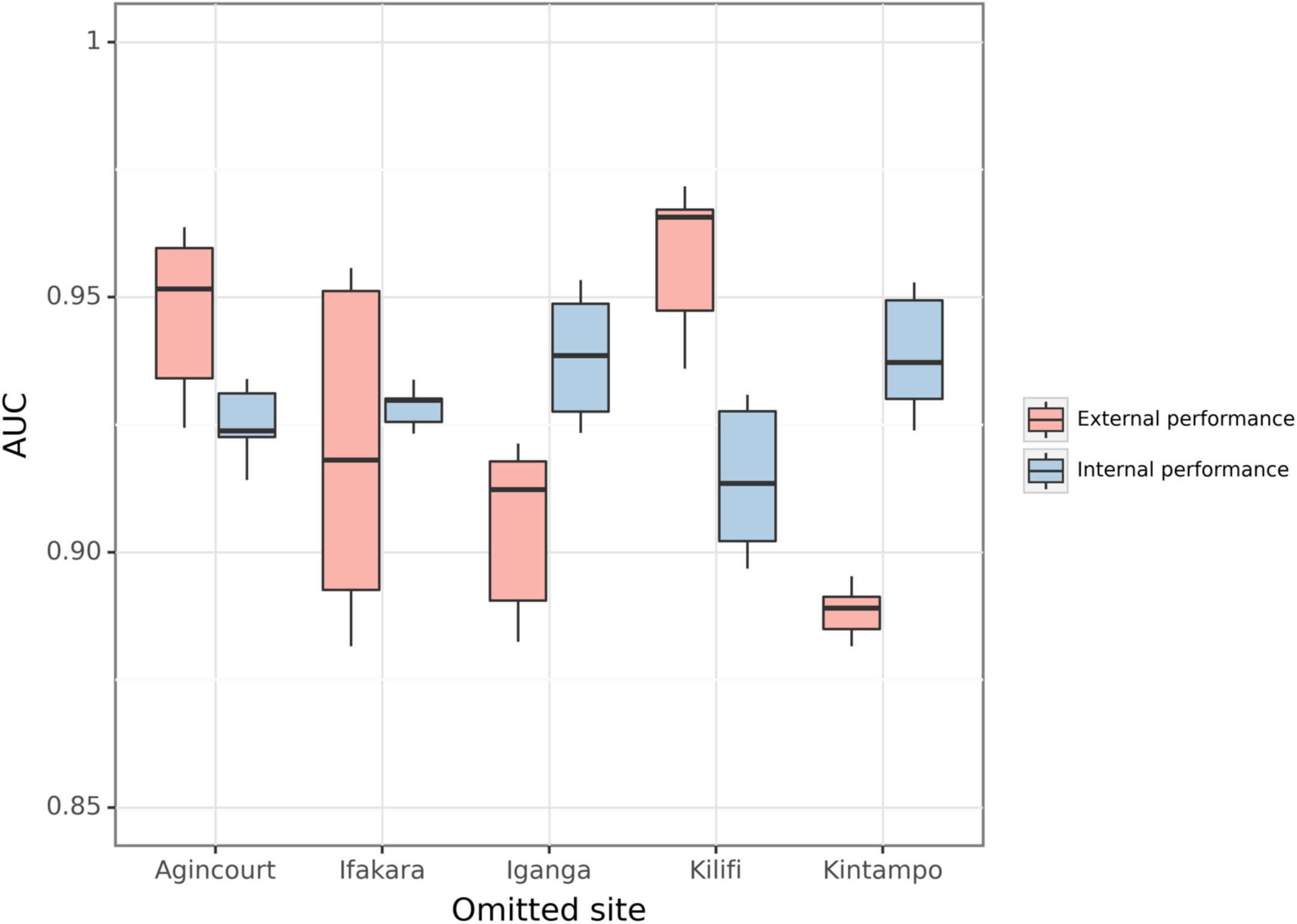
Performance of LOSO models. Boxplot of AUC values resulting from the testing, on each site in turn, of models trained on all but one site. Both internal and external performance is shown. Internal performance was higher in 3 sites (internal median 0.93, external median 0.89). In the other two, AUC values displayed a larger range (internal median 0.92, range 0.04; external median 0.96, range 0.05). AUC = area under receiver operating curve. Internal performance = performance on sites included in training data. External performance = performance on the site that was not included in training data. Central line is median; lower edge of the box indicates first quartile, upper edge of the box indicates third quartile; points are extreme performance scores.

**Figure 5.**
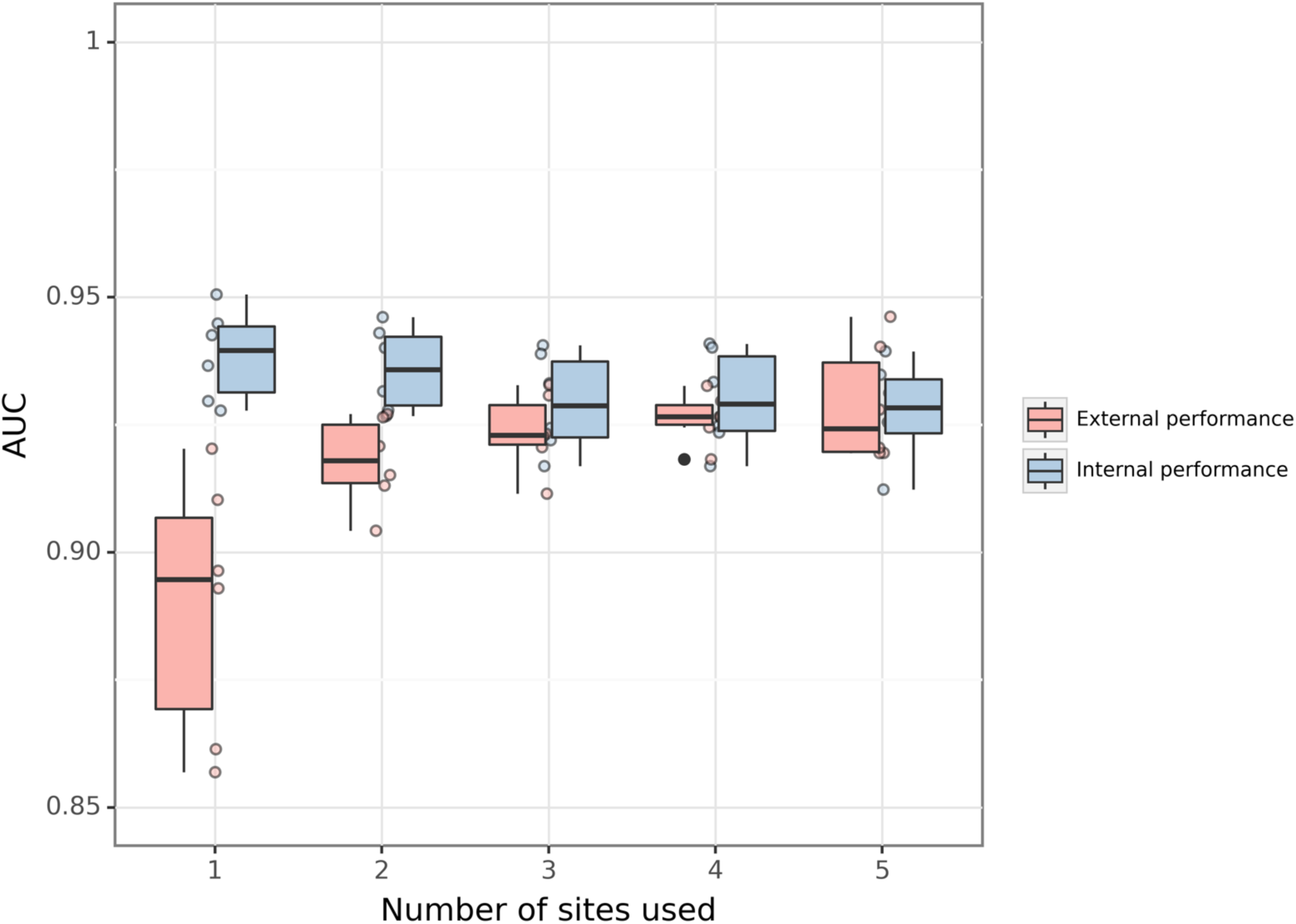
Performance of incremental models. Boxplot (with scatter graph) showing the change in AUC as the number of sites included in training is increased. Both internal and external performance is shown. As sites were added, internal performance worsened and external performance improved. At the stage when only one site was included in the training data, all external performances were lower than all the internal performances. At the final stage, the two measures converged to a small final difference, and all the data points from the external validation were higher than all the internal. AUC = area under receiver operating curve. Internal performance = performance on sites included in training data. External performance = performance on the site that was not included in training data. Central line is median; lower edge of the box indicates first quartile, upper edge of the box indicates third quartile; scatter points are individual performance scores.

Agincourt (internal median 0.924, IQR 0.008; external median 0.95, IQR 0.03; p<0.05) and Kilifi (internal median 0.91, IQR 0.025; external median 0.97, IQR 0.02; p<0.01) had higher external performance, and these values displayed a larger range (Agincourt: p=0.08; Kilifi: p=0.65).

Patterns emerge when comparing the one-site models with the LOSO models. For instance, both model types showed that Agincourt and Kilifi manifested a wider range of external performance scores than internal performance scores, with the inverse relationship evident at the other sites. Agincourt and Kilifi demonstrated superior external median performance in the LOSO model relative to their internal performance, and a trend contradicted at the remaining sites. Kintampo consistently presented the lowest external variability in LOSO and one-site models, further underscoring the site-specific patterns inherent in the performance of these diagnostic tools.

## Discussion

We demonstrate that deploying an epilepsy diagnostic model outside the cultural and geographical region in which it was developed can result in highly unpredictable, frequently sub-optimal outcomes.

As with other ML models, the generalisability of clinical epilepsy diagnostic tools is inherently constrained. Extrapolating these models beyond their original regional parameters necessitates a trade-off between internal and external performance. Performance scores within a given area consistently exhibit lower variances and higher medians than those obtained from cross-site applications. This is perhaps especially problematic given the volume of ML-driven diagnostic models for epilepsy that have been developed using single regions (25)(26). Incorporating data from a greater number of sites into the model’s training set can help mitigate the risk of ML models making erroneous assumptions and thereby offering incorrect diagnoses. Whilst this approach engenders a model with enhanced robustness and improved external performance, it concomitantly decreases internal performance. This occurs as the model broadens its applicability while becoming less tailored to specific sites. While analysis of generalizability has been performed elsewhere (10), this study presents a novel and important contribution to the field of epilepsy diagnosis.

The weight of each predictor for ACE, and the optimal thresholds for a classification vary significantly between sites. At the extreme, a particular symptom might correlate positively with ACE at one site and negatively at another. These disparities may result from the variance in reporting of epilepsy-related symptoms across geographies.

Data underscore the importance of thoughtfully calibrating diagnostic procedures to the unique specificities of each geographic locale. The observed variability in thresholds also highlights that a simplistic transposition of one site’s threshold for ACE classification to another may lead to an unpredictable number of misdiagnoses, most concerningly false negatives, where an individual remains undiagnosed and cannot access treatment. The findings emphasise the necessity of adjusting model parameters to ensure their suitability for application in varying settings.

A variety of explanations may account for differences in performance between the sites. One reason may be that symptoms may be reported differently by those with epilepsy or their carers, depending on the cultural and clinical context. These cannot necessarily be predicted and accounted for and may differ between sites. For example, appearing dazed during episodes was the single most prevalent symptom reported by individuals with seizures in Iganga and one of the least prevalent in Ifakara (Supplementary Figure 3).

Based on the results of this study, we suggest that some degree of site-specific validation is essential before a predictive model is deployed in practice. While complete re-training of all model parameters on local data would be ideal and result in optimal performance, this is not always feasible. Studies have shown that merely changing the intercept of a logistic regression model may help recontextualise a model for new settings (27). It may suffice to adjust only a few parameters.

As performance is continually assessed, iterative updating of the model throughout deployment can help fine-tune the model further. Such a model would generally need a threshold to determine who is deemed a positive case, although, as shown, optimal thresholds vary between sites. Validating the threshold may also effectively update the model to perform well in a new setting – we saw a mean change in accuracy of 10% following a threshold change.

### Limitations

This work is limited by the choice of dataset and its contents. The data were acquired from five distinct sites from five different sub-Saharan African countries. We cannot necessarily draw conclusions about, for example, training a model on data from a specific area and deploying it in a nearby location in the same country with similar demographics. The dataset also featured limited accounting of seizure type. The results could be clouded by, for example, the effects of non-convulsive seizure phenomena, which we could not account for.

We attempted to minimise this inherent heterogeneity by separating the generalised seizures out into a separate dataset. Generalised convulsive seizures were selected as they demonstrate a higher degree of homogeneity in their clinical presentation than focal seizures (9). Further work should explore focal seizures to identify better how ML models may also help in their diagnosis.

There also may have been data collection and data entry issues: self-reporting may be influenced by linguistic differences in how questions were asked, cultural differences in how chronic conditions are perceived, or who asked the questions and how. The clinicians making diagnoses or the workers performing data entry may also vary in reliability. While the tools and questions were adapted to the local context and training was standardised before data collection and monitored during (11), human factors may still play some role.

### Conclusions

We demonstrate that, when developing models for epilepsy diagnosis, data collected from one site cannot naïvely be used as representative for all other sites where a model could be deployed. Given the needs of LMICs, ML models can be leveraged to significant impact. Nonetheless, careful interrogation and a dedication to rooting tools in the setting of their use must be ensured to avoid such models associating with inadvertent harm, including missed diagnoses resulting in delayed initiation of care. Whilst the present analysis has focused on convulsive epilepsy, similar arguments are likely applicable across other seizure types and disorders of mind-brain health more broadly.

## Data Availability

The authors do not have permission to publicly share the data used. Given the multisite origin of these data, requests for data will require approval from the clinical sites and partner institutions requests can be made to the corresponding author. Queries regarding access to the data in the event the corresponding author is no longer available for contact should be addressed to Charles R Newton.

## Collaborators

- Study of Epidemiology of Epilepsy in Demographic Sites (SEEDS)
- Oxford Martin Programme on Global Epilepsy https://www.oxfordmartin.ox.ac.uk/global-epilepsy/)
- Epilepsy Pathway Innovation in Africa (EPInA, https://epina.web.ox.ac.uk/)

---

|  | Study site |  |  |  |  |  |  |
| --- | --- | --- | --- | --- | --- | --- | --- |
| Convulsive epilepsy predictor | Agincourt | Ifakara | Iganga | Kilifi | Kintampo | Median | p-value |
| Do you experience stomach ache before these episodes? | 0.7 | 1.6 | 2.1 | 2.3 | 1.1 | 1.6 | 0.001 |
| Do you see odd things (e.g. flashes or bright lights) before these episodes occur? | 1.4 | 1.9 | 1.3 | 1 | 0.8 | 1.3 | <0.001 |
| Do you think anything brings on these episodes? | -0.5 | -4.3 | -2.8 | -4.3 | -2 | -2.8 | <0.001 |
| During these episodes, do you lose contact with your surroundings? | 1.5 | 2.3 | 0.4 | 1.2 | 2.6 | 1.5 | <0.001 |
| During these episodes, does your body stiffen? | 2.8 | 2.8 | 0.4 | -0.1 | 1.7 | 1.7 | <0.001 |
| During these episodes, have you ever bitten your tongue? | 0.9 | 2.5 | 0.9 | 0.8 | 2.5 | 0.9 | <0.001 |
| Has anyone told you that you appear dazed during these episodes? | 0.8 | 1 | 3.2 | 1.3 | 3 | 1.3 | <0.001 |
| Have you ever wet yourself during these episodes? | 1.5 | 1.9 | 1.6 | 1.3 | 2.2 | 1.6 | <0.001 |

## Supplementary figures

**Supplementary Figure 1.**
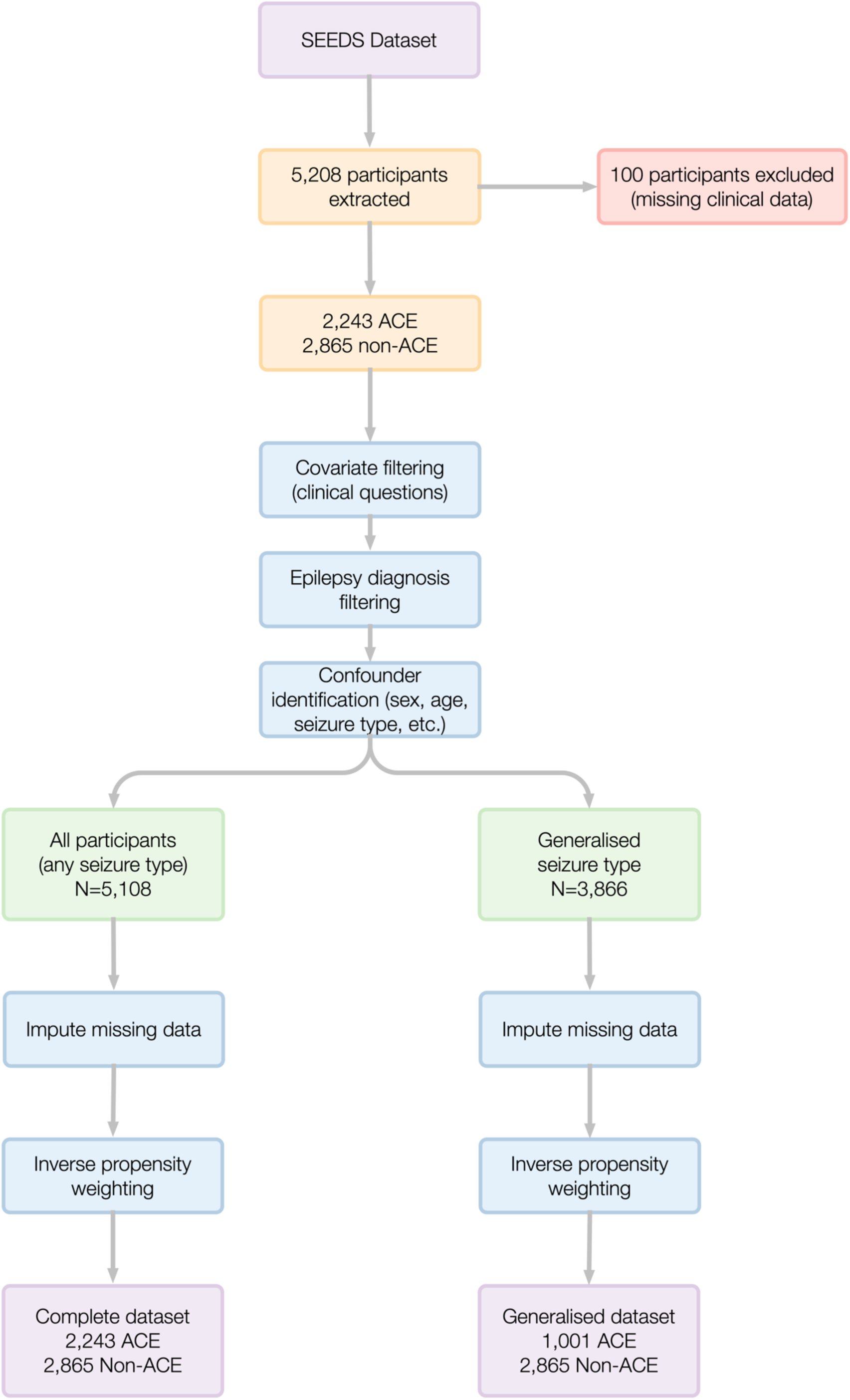
Data preparation. Flowchart showing the process of data preparation (see Table 1).

**Supplementary Table 1.**
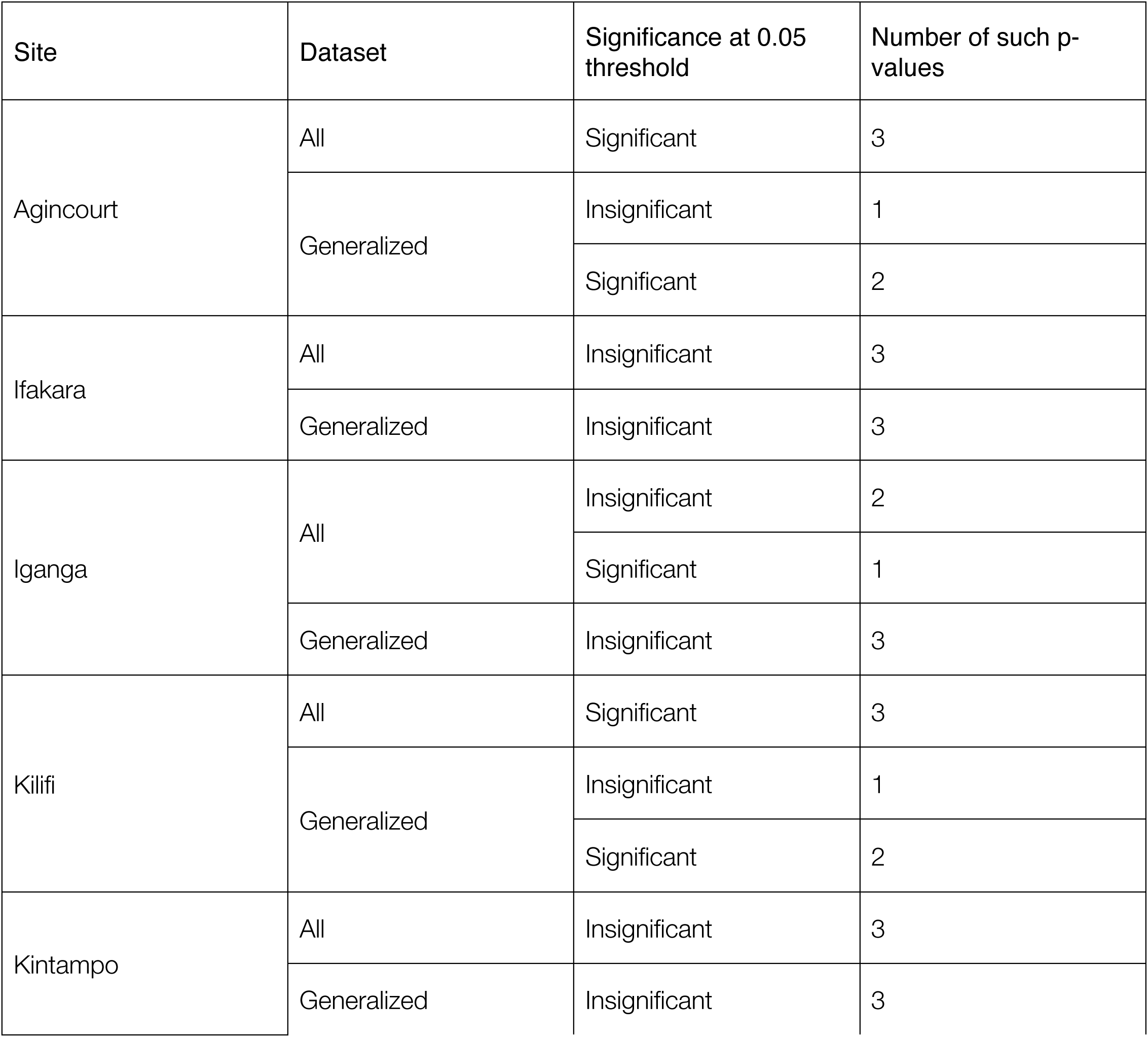
Kolmogorov-Smirnov test p-values. Table summarizing the p-values of the statistical tests of the performances resulting from the testing of each one-site model on each of the other sites in turn. The two samples in each test were the internal performances for that site (from 5-fold cross validation) and the performances of that model on each other site. This was done for each possible combination of the three model types (Logistic Regression, SVM using a linear kernel, and Naive Bayes assuming a Bernoulli distribution) and the two datasets (the whole dataset, and that with the positive cases limited to participants with generalised epilepsy). Thus, there are 6 values for each site. The 2-sample Kolmogorov-Smirnov test has the null hypothesis that the two samples are drawn from the same distribution. Statistical insignificance is taken as insufficient evidence to reject this. There were 7 significant tests for the whole dataset and 4 for the generalised seizure dataset. Agincourt and Kilifi each had 5 significant tests, while the other 3 sites each had at least 5 insignificant tests.

**Supplementary Figure 2.**
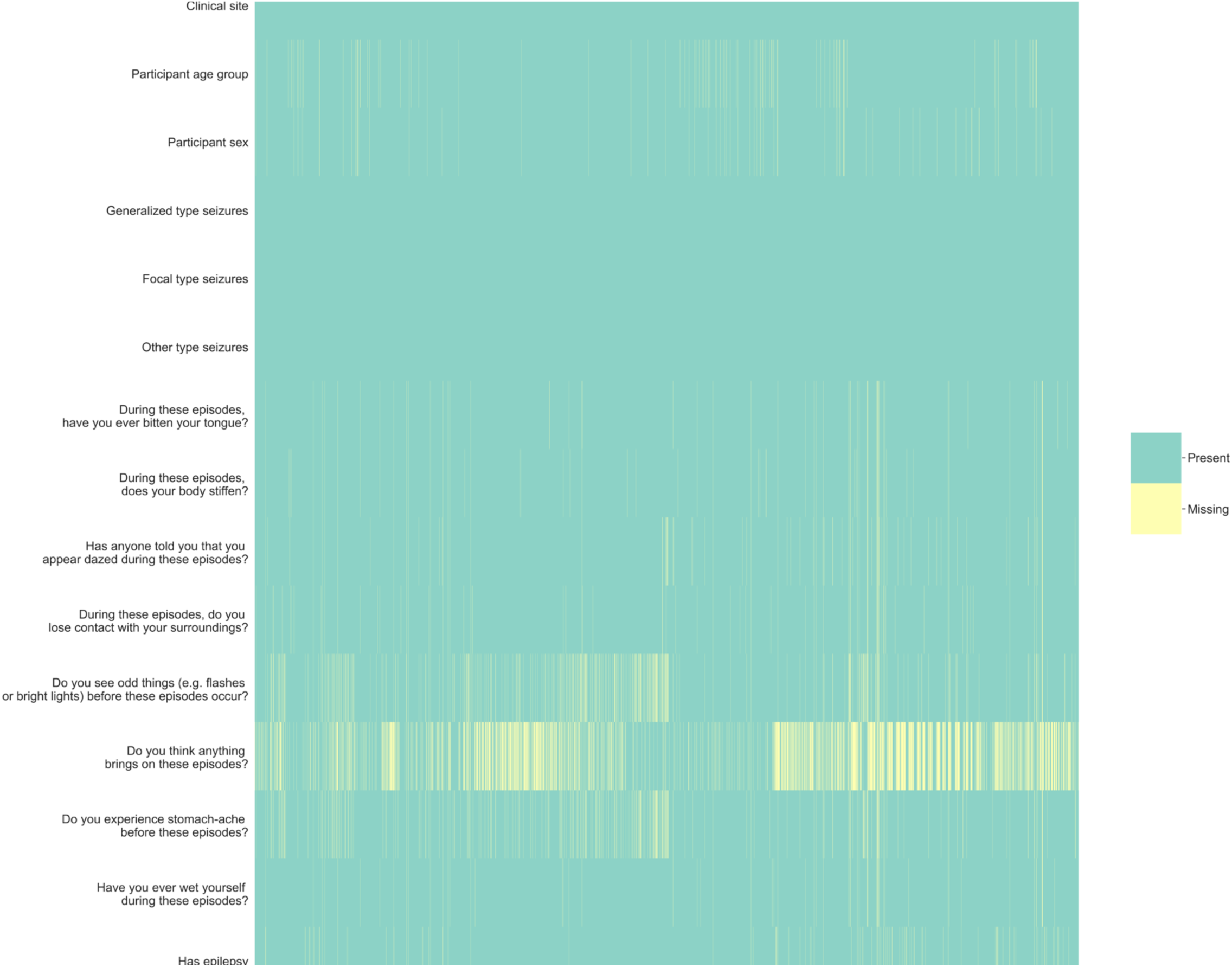
Missingness in the raw data. Heatmap showing where there was unexplained missingness in the data before cleaning. Missing values are shown in yellow, others in green. The data are sorted according to assessment date, in ascending order from earliest to latest.

**Supplementary Figure 3.**
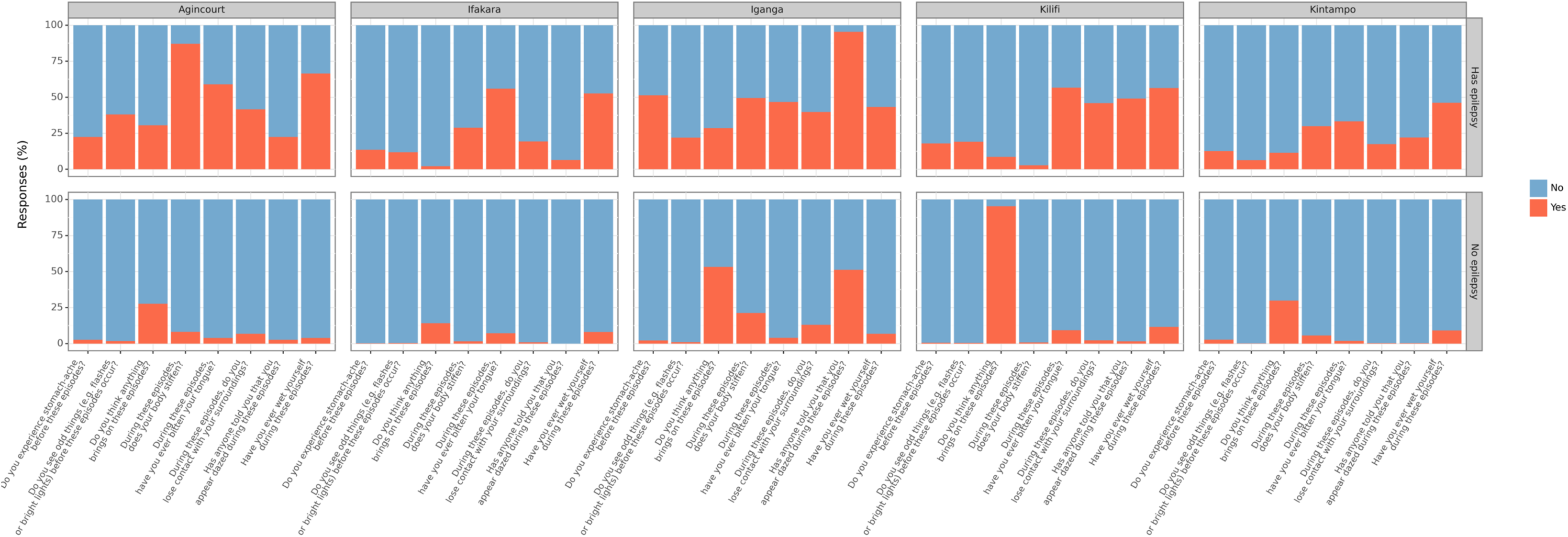
Covariate values per site. Stacked bar chart showing the percentage of yes/no answers to the covariate questions. Values taken from the cleaned data, split by site and epilepsy diagnostic class.

**Supplementary Figure 4.**
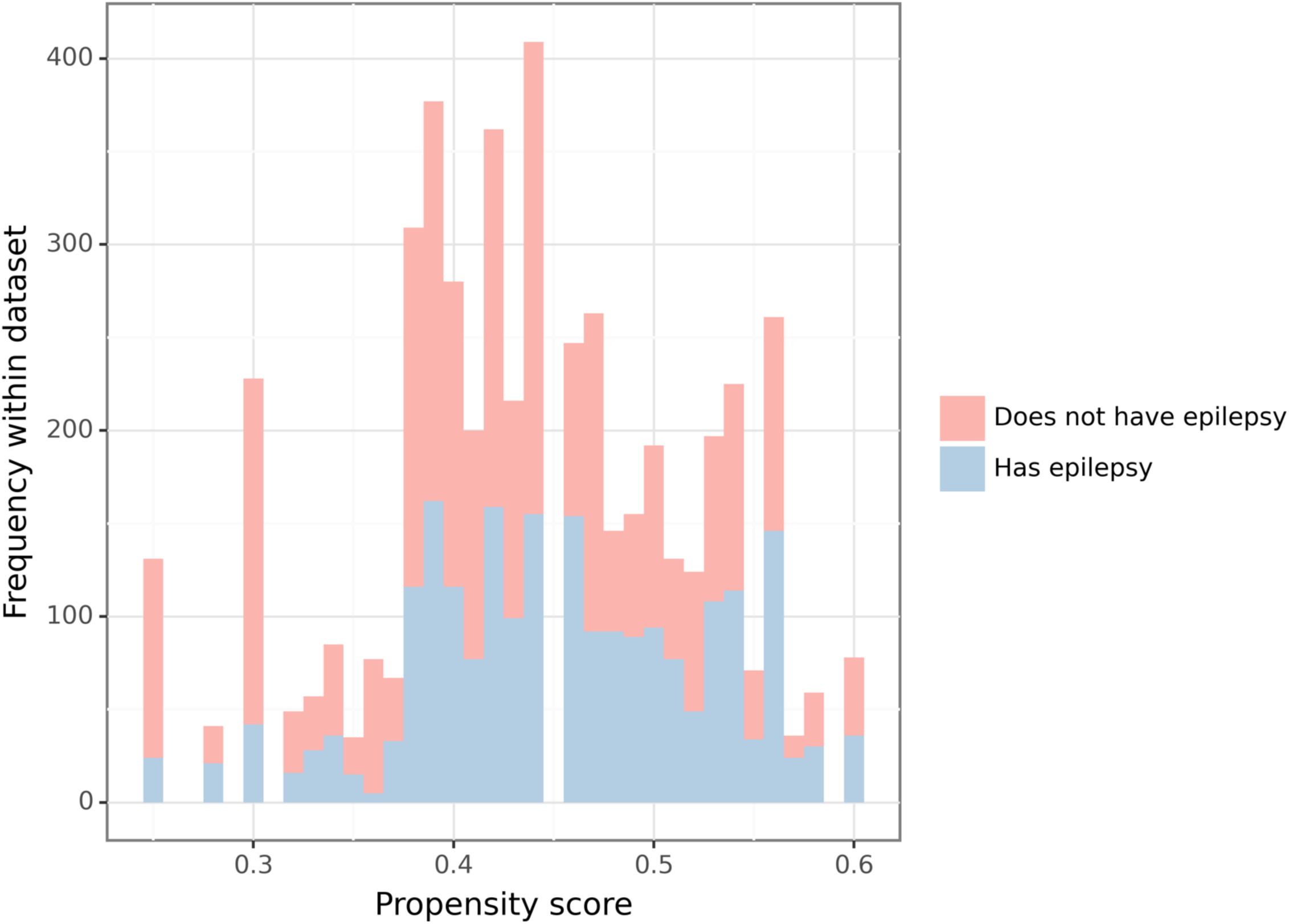
Histogram showing propensity scores as calculated from the processed data, coloured by whether there was a diagnosis of epilepsy (orange) or not (blue). There is a complete overlap – the range of scores for the two diagnostic classes overlaps completely.

